# Genomics-Informed Drug Repurposing Strategy Identifies Novel Therapeutic Targets for Metabolic Dysfunction-Associated Steatotic Liver Disease

**DOI:** 10.1101/2025.02.18.25321035

**Authors:** Hannah M. Seagle, Alexis T. Akerele, Joseph A. DeCorte, Jacklyn N. Hellwege, Joseph H. Breeyear, Jeewoo Kim, Michael Levin, Samuel Khodurksy, Adam Bress, Kyung Lee, Jens Meiler, Dipender Gill, Jennifer S. Lee, Kent Heberer, Donald R. Miller, Peter Reaven, Kyong-Mi Chang, Julie A. Lynch, Nikhil K. Khankari, Megan M. Shuey, Todd L. Edwards, Marijana Vujkovic

**Author notes:** Contributed equally as senior author (Megan M. Shuey, Todd L. Edwards, Marijana Vujkovic).

## Abstract

Identification of drug-repurposing targets with genetic and biological support is an economically and temporally efficient strategy for improving treatment of diseases. We employed a cross-disciplinary approach to identify potential treatments for metabolic dysfunction associated steatotic liver disease (MASLD) using humans as a model organism. We identified 212 putative causal genes associated with MASLD using data from a large multi-ancestry genetic association study, of which 158 (74.5%) are novel. From this set we identified 57 genes that encode for druggable protein targets, and where the effects of increasing genetically predicted gene expression on MASLD risk align with the function of that drug on the protein target. These potential targets were then evaluated for evidence of efficacy using Mendelian randomization, pathway analysis, and protein structural modeling. Using these approaches, we present compelling evidence to suggest activation of *FADS1* by icosopent ethyl as well as *S1PR2* by fingolimod could be promising therapeutic strategies for MASLD.

## Introduction

The high prevalence of cardiometabolic diseases reflects, in part, the consequence of the global obesity epidemic. In the past five decades, the incidence of obesity has increased threefold, with the anticipated global prevalence of obesity and overweight individuals expected to reach 51% by 2035.^1^ In the United States, the prevalence is anticipated to be 78% by 2030^1,2^, with a collateral rise in associated disorders such as metabolic dysfunction-associated steatotic liver disease (MASLD). This increase has important implications for MASLD, formerly known as non-alcoholic fatty liver disease, as there are limited treatment options. Resmetirom, the first and only approved treatment of metabolic dysfunction associated steatohepatitis (MASH), an advanced form of MASLD, received United States Food and Drug Administration approval in March 2024.^3,4^

Prior to the approval of Resmitirom, most treatment and management strategies for MASLD focused on lifestyle interventions. Initial drug repurposing efforts, or finding new uses for existing medications, have been primarily focused on trials of diabetes treatments such as pioglitazone, glucagon-like peptide 1 receptor (GLP1R) agonists, or sodium-glucose transporter 2 (SGLT2) inhibitors. The rationale for this approach is two-fold: first, hyperinsulinemia and insulin resistance play an important role in the pathogenesis of MASLD;^5–7^ second, these treatments have demonstrated efficacy in reducing intra-organ fat or body weight and lipid levels^8–12^, which are two significant risk factors for MASLD. If successful, these treatments would benefit the ∼18.2 million Americans diagnosed with type 2 diabetes and MASLD.^13,14^.However, not all patients with MASLD have diabetes and MASLD may predate diabetes onset. Thus, focusing exclusively on diabetes-specific medications may not address the needs of all MASLD patients.

It is therefore essential to expand our efforts, including strategies such as drug repurposing to identify effective therapeutics. As many drugs modulate the expression or function of additional molecules beyond their original targets, assessing their potential for repurposing is an attractive strategy for identifying new uses for approved or investigational drugs that are outside the clinical indication of the original medication. Historically, drug repurposing was the result of chance discovery of off-target effects in routine clinical care. An example is the repurposing of the anti-hypertensive medication, sildenafil citrate, for erectile dysfunction.^15^ The economic incentives for drug repurposing are substantial; the average cost to market a repurposed drug is $300 million, which is a fraction of the $2-3 billion typically required for bringing a new drug target to market.^16,17^

Human genetic studies are becoming increasingly important in drug discovery. A notable example is the identification of monoclonal antibodies that target interleukin-23 to treat Crohn’s disease^18,19^ and PCSK9 inhibitors for reducing LDL cholesterol^20^ from genome-wide association studies (GWAS). We, and others, have demonstrated how genetic discovery can nominate repurposing drug targets considering transcription as well as validation using Mendelian randomization (MR).^21,22^ Further, modern computational approaches to protein modeling and drug docking now allow one to create high-fidelity *in silico* models of drug-protein interactions.^23–25^ These models provide additional support for drug repurposing efforts through improving efficiency by identifying the results of genetic studies that have a strong structural rationale for binding to a protein of interest.

In this paper, we present a genomics-informed drug repurposing prioritization strategy to identify existing drugs for potential treatment of MASLD using the human as a model organism. Using a large multi-ancestry GWAS of MASLD, we performed a transcriptome wide association study (TWAS) and colocalization analysis to identify gene expression signatures associated with MASLD risk. We then used mapping strategies and examined direction of effect to identify MASLD genes targeted by existing drugs (drug-gene pairs). Using MR, we tested whether a causal relationship exists between the drug-gene target and MASLD risk. We then utilized protein-ligand modeling for the significant causal genes to confirm drug targets and nominate additional potential therapeutics. Finally, we used the significant signals from the GWAS analysis to identify regulatory pathways and networks and compared the primary drivers of these mechanisms to those identified in the previous approaches to further support our target prioritization. By utilizing this cross-disciplinary approach, we identified three and prioritized two potential repurposing targets for MASLD treatment.

## Methods

### MASLD Phenotype Definition and GWAS

The overall study design is presented in Figure 1. We utilized previously-published summary statistics from our recent multi-ancestry independent GWAS for MASLD including 90,408 MASLD cases and 128,187 controls in the Million Veteran Program (MVP).^26^ In that study, MASLD cases were defined as: (1) elevated alanine aminotransferase (ALT) > 40 U liter^−1^ for males or >30 U liter^−1^ for females during at least two time points at least 6 months apart within a 2-year window at any point prior to enrolment, and (2) exclusion of other causes of liver disease, chronic liver diseases or systemic conditions and/or alcohol use disorders. Control subjects had normal ALT levels and no apparent causes of liver disease or alcohol use disorder or related conditions. This previously published multi-ancestry meta-analysis identified 77 independent variants associated with MASLD, of which 17 variants were replicated in an external histologic cohort in which MASLD was defined using liver biopsies and an independent radiologic cohort where hepatic triglyceride content was determined using reports from computed Tomography (CT/) and magnetic resonance imaging (MRI) data of the liver and spleen.

**Figure 1.**
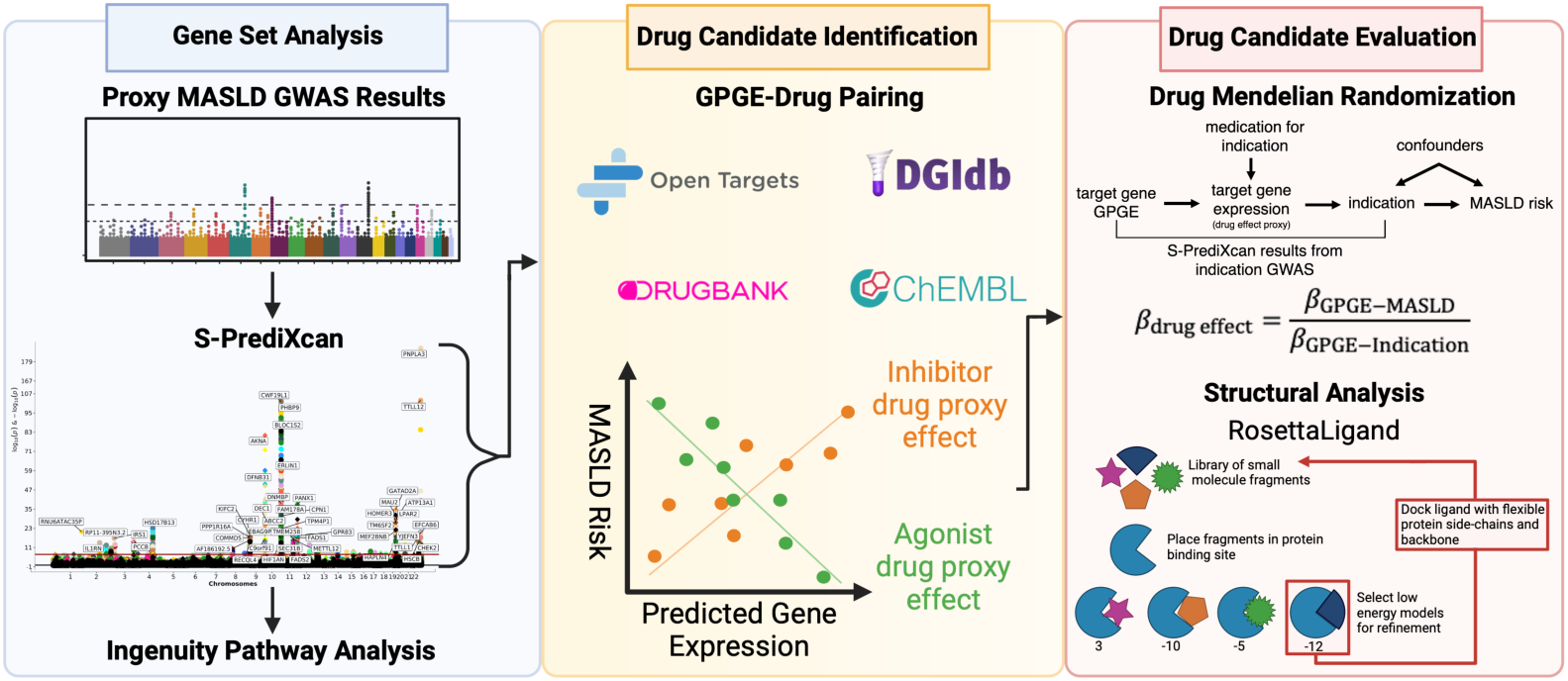
Schematic overview of analytical framework.

### Gene Set Analysis

Given that most GWAS loci do not directly cause disease and are infrequently located within a protein-coding region of the genome, mapping MASLD-associated variants to putative causal genes is important to ensure the correct druggable target is selected for downstream analysis. For the prioritization of putative candidate genes associated with MASLD risk, we previously accumulated eight lines of human genetic evidence. This included coding variant linkage analysis, colocalization analysis with gene expression, splicing expression, histone, and chromatin accessibility data, liver-specific enrichment analysis, and protein-protein interaction network analysis.^26^ For each MASLD-associated locus, the gene that was most frequently identified as a causal gene in these eight experiments was selected as the putative causal gene. In addition to these approaches, we further identified putative effector genes through colocalization analysis with protein levels as well as genetically predicted gene expression analysis, as described in the following sections. The resulting updated list of effector genes was subsequently mapped to known drug targets, and the final set of drug-gene pairs were then taken forward to drug candidate evaluation.

### Identifying MASLD genes using MR and protein level data

Our first MR was performed to evaluate the effects of circulating proteins on risk of MASLD. Genetic instruments were constructed from genetic variants associated with circulating proteins (p < 5×10^-8^), identified from the deCODE GWAS of 4,907 circulating proteins measured using an aptamer-based method among 35,559 Icelanders.^27^ Uncorrelated cis-acting genetic variants (r^2^ < 0.01) located within +/-250kb of the gene encoding each target protein were used to construct genetic instruments. Corresponding effects of each protein-associated variant on MASLD were obtained from the multi-ancestry MVP GWAS.^26^ MR was performed using inverse variance weighting, accounting for residual linkage disequilibrium using the 1000 Genomes Phase 3 EUR reference panel using the *MendelianRandomization* package in R.^28^ When a single variant within this window was present, MR was performed using the Wald ratio. The false discovery rate was controlled at 5% to account for multiple comparisons.

For circulating proteins significantly associated with MASLD in the MR study, colocalization^29^ was performed to provide complementary genetic evidence to support a shared genetic signal influencing both circulating protein levels and risk of MASLD.^30^ We considered the presence of colocalization (posterior probability that both protein levels and MASLD risk are associated at the same causal variant) greater than 70% as genetic support for colocalization.

### Predicting gene expression profiles of MASLD genes

Given the limited availability of available gene expression data in diseased tissues, we aimed to predict the expression levels of MASLD-associated genes in 48 healthy tissues as a function of germline genotypes. Advantages of using all tissues as opposed to liver tissue alone include increased power for discovery and shared eQTLs across tissues. S-PrediXcan was used to estimate genetically predicted gene expression (GPGE) using the MASLD GWAS summary statistics. Details regarding S-PrediXcan^31^ have been previously described. Briefly, S-PrediXcan integrates gene expression prediction models from the Gene-Tissue Expression Project (GTEx v7)^32^, covariances sourced from a reference dataset, and variant-specific effect estimates alongside standard errors extracted from GWAS summary statistics. This information enables the prediction of tissue-specific alterations for a given trait per standard deviation (SD) increase in gene expression levels. The Bonferroni-corrected significance threshold was 2.03×10^-7^.

Colocalization^29^ was used to identify shared variants significantly associated with MASLD and gene expression at a particular locus. The expression quantitative trait loci (eQTL) summary statistics corresponding to the gene-expression references utilized in the GPGE analysis and the common variants from the multi-ancestry meta-analyses, restricted to variants included in the S-PrediXcan models, were included as input for colocalization analysis. Statistically significant S-PrediXcan results with a corresponding posterior probability for the fourth alternative hypothesis greater than 80% were considered to have strong evidence for colocalization.

### Gene pathway analysis

Ingenuity Pathway Analysis (IPA) (Qiagen) software was used to identify enriched pathways and networks from all significant genes (P < 2.03×10^-7^) associated with MASLD in the S-PrediXcan analysis described above. IPA software analyses have been previously described. All analyses were ordered by enrichment p-value, including canonical signaling, diseases and functions, and regulatory networks.^33^

### Drug Candidate Identification

We adapted a previously developed genetically informed drug-repurposing pipeline^21,34^ for MASLD drug discovery. All MASLD associated effector genes were mapped to multiple drug targets using publicly available well-curated databases: OpenTargets^35,36^ and Drug Gene Interaction Database (DGIdb).^37^ If identified as having a known drug target in these databases, genes were included as a drug-gene pair for further evaluation. These drug-gene pairs were further refined by direction of GPGE and drug effect. For example, inhibitors (or blockers) were identified for increased gene expression associated with increased MASLD risk, and vice versa. This final number of drug-gene pairs with directional concordance of effect were considered as potential experimental medications for MASLD management and were taken forward in our *in silico* drug repurposing analysis. Duplicate medications and those with a primary indication for cancer treatment were excluded due to severe adverse event profiles. All medications under consideration for treating MASLD were cross-referenced with data from NCBI LiverTox^38^ and the FDA DILIRank^39^ drug list to extract information on safety profile and known hepatotoxicity.^39^

### Drug Candidate Evaluation

Our *in silico* drug repurposing evaluation using MR was based upon the list of identified drug-gene pairs with concordance of direction of effect. First, we collected GWAS summary statistics for the primary indications for these drugs and generated separate GPGE, using S-PrediXcan. These GPGE summary statistics were then used as instrumental variables to proxy therapeutic targets in the MR analysis to identify potential off-target effects for reducing MASLD risk^21,34^ (Supplemental Table 1).

Briefly, two-sample MR was conducted using GPGE summary statistics (derived from S-PrediXcan) for each selected drug-gene target to proxy current medications. Tissue-specific GPGE summary statistics, for each proxied drug-gene target, were estimated using S-PrediXcan (as described above) and were used in tandem with the medication GPGE to conduct a two-sample MR. This analysis utilized tissue-specific GPGE summary statistics that met the p-value threshold of 0.05 as the instrumental variable and was conducted using the “TwoSampleMR” R package.^40^ All tissue-specific GPGE at a specific gene target were harmonized to represent either an increase or decrease in target gene expression according to the mechanism of drug action. For example, for inhibitors, the GPGE effects at that gene target represented changes in the primary indication associated with one SD decrease in gene expression. After harmonization, the GPGE effects at each drug target were summarized across all statistically significant tissues using random-effects meta-analysis (Supplemental Table 1) to estimate predicted effects of the proxied drugs (via specific gene) on genetically predicted changes in the primary indication.

We then performed an inverse-variance weighted (IVW) MR for instruments with > 1 GPGE. For gene targets with only 1 GPGE, MR effects were estimated using the Wald ratio estimator. Sensitivity analyses were performed to assess the robustness of our MR results. To evaluate bias due to directional pleiotropy at the tissue level, MR Egger^41^ was conducted. Statistically significant regression intercepts (P < 0.05) were an indication of directional pleiotropy. Additionally, the heterogeneity of tissue-specific MR effects was evaluated using the Q statistic. For those indications for which MR effects were heterogenous, we estimated inflated standard errors and corresponding confidence intervals using random-effects IVW MR.^42^ Genes with significant MR results were carried forward for structural analysis.

### Modeling of protein-drug complexes

All complexes were prepared in Rosetta 3.13 with RosettaLigand.^43^ Rosetta parameters for each ligand were created using conformers generated in the Biology and Chemistry Library (BCL)^44^, an open-source cheminformatics platform. All relevant cofactors were included during ligand docking. XML scripts for docking will be provided upon request.

### Protein preparation for molecular docking

When an experimental structure (e.g., crystallography or cryo-electron microscopy) existed of a protein of interest bound to a ligand/ligands, these structures were prioritized for binding-site selection. For proteins with only an apo-state experimental model, binding sites were identified by consensus modeling using DiffDock and AlphaFold3.^45^ Missing loops in experimental structures were modeled by generalized kinematic closure in Rosetta. For proteins with no published experimental structures, structural models were created in AlphaFold3, then refined through Rosetta’s relax and minimization protocols prior to consensus binding-site identification as above.

### Molecular dynamics simulations

Molecular dynamics (MD) simulations were performed in AMBER^46^ using the ff19SB force field for proteins; Optimal Point Charge water model for solvent molecules; general AMBER force field for small-molecule ligands; and, when lipid membranes were used, the lipid21 force field for lipids. The system was prepared through minimization, heating, and equilibration steps, followed by production runs, all under NpT (isothermal-isobaric ensemble) ensemble conditions. For soluble proteins, minimization was carried out first for the solvent, then for the entire system; heating occurred gradually from 0K to 310K; and production simulations were conducted unrestrained at 310K under NpT conditions. SHAKE was used during production runs but not minimization, and hydrogen mass repartitioning^47^ was used throughout, allowing 4fs timesteps using a Langevin integrator during production simulation. Integral membrane proteins were embedded into a pre-equilibrated lipid bilayers of appropriate composition using the CHARMM-GUI.^48^ System minimization, heating, equilibration, and production were performed as suggested by CHARMM.

## Results

### Contextualizing Genetic Signals

The original multi-ancestry meta-analysis identified 77 independent variants associated with MASLD, which prioritized a total of 81 putative causal genes for these loci.^26^ In the current study, we identified 3 additional genes using protein level MR and colocalization analysis, and 160 genes using GPGE analysis across 48 tissue types, of which 29 were overlapping with the GWAS-based gene set of 81 causal genes (Supplemental Table 2A/B). Combined, these analyses yielded a total of 212 unique putative causal genes for MASLD. Of these, 158 genes (74.5%) have not been previously associated with MASLD or liver enzyme levels in the GWAS Catalog or OpenTargets at the time of search. We identified a total of 412 statistically significant (P = 2.03×10^-7^) gene-tissue pairs associated with MASLD that also had a high posterior probability (PP.H4 > 0.8) of colocalization (Supplemental Table 3). Of these pairs, 23 included a gene with significant MR results, where *S1PR2* expression was strongly associated in liver tissue (P = 4.63×10^-9^, PP.H4 = 0.90).

The results of the pathway analysis, including canonical signaling, diseases and functions, and regulatory networks, are presented in Supplemental Table 4. The top two canonical pathways included FXR/RXR activation and LXR/RXR activation (P = 6.81×10^-12^ and 9.93×10^-8^, respectively, Supplemental Table 4). The prominence of rexinoid activity further supports the role of *PPARG* observed above and may suggest a broader disease mechanism. Other significant findings included gastrointestinal, hepatic system, and metabolic disease, as well as lipid metabolism and increased levels of potassium, alkaline phosphatase (ALP), ALT, and albumin (Supplemental Table 4). Taken together, these results align with our expectations for metabolic and liver-related conditions and highlight the involvement of genetic signals in these metabolic and liver disease pathways.

### Drug Candidate Identification

Building on a previously developed genetically informed drug-repurposing pipeline, we identified a total of 57 drugs with MASLD repurposing potential. These drugs targeted 13 unique genes, 6.67% of the total genes identified from the TWASs and three proteins, 7.3% of the total unique proteins identified from the protein Quantitative Trait Loci (pQTL) MR analysis. Of 57 drugs, 11 were local anesthetics (mapped to *SCN2A*) and 33 are listed in either the LiverTox^38^ or FDA DILI database^39^, of which five are flagged as class A (well-known cause of clinically apparent liver injury), three as class B (likely rare cause of clinically apparent liver injury), and 10 as class C (probable cause of clinically apparent liver injury) in NIH NIDDK LiverTox Database, whereas FDA classified eight of the 57 drugs as having most concern for drug-induced liver injury with confirmed causal evidence linking a drug to liver injury.

### Drug Candidate Evaluation

Results of the MR analyses proxying the effects of medications on MASLD risk are presented in Table 1. Across the five significant indications, six gene targets were used as proxies for the therapeutic action of repurposing existing drugs for treatment for MASLD. These genes mapped to five drugs, of which three had a significant effect on MASLD risk, namely icosapent ethyl, pioglitazone hydrochloride, and fingolimod which are proxied by *FADS1/2, PPARG, and S1PR2* respectively. Increased expression of *FADS1* and *S1PR2*, proxies for icosapent ethyl and fingolimod, respectively, showed significant potential to lower MASLD risk (*FADS1:* IVW odds ratio (95% CI) = 0.28 (0.26–0.31), P = 6.8×10^-160^; and *S1PR2:* IVW odds ratio (95% CI) = 0.91 (0.86–0.97), P = 4.0×10^-4^ Table 1). Conversely, upregulation of *PPARG* expression, a proxy for pioglitazone hydrochloride, a drug that is used to treat type 2 diabetes, was associated with an increased MASLD risk (P = 1.2×10^-14^, IVW odds ratio (95% CI) = 1.93 (1.62–2.31), Table 1).

**Table 1.**
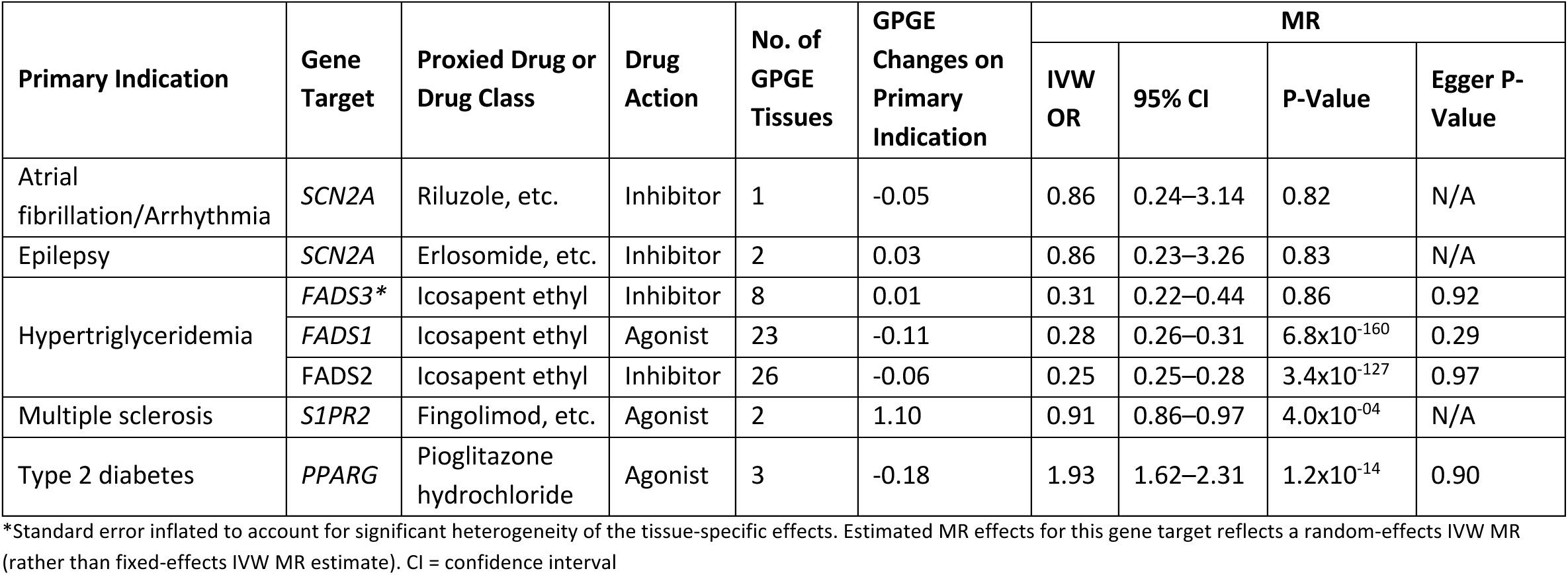
Changes in MASLD risk per standard deviation change in GPGE, as a proxy for the therapeutic action of repurposing existing drugs or treatment of MASLD via fixed-effects IVW MR.

### In silico modeling of protein-drug complexes

#### PPARG

We identified pioglitazone hydrochloride, a PPARG agonist, within the thiazolidinedione (TZD) class of drugs, as a potential contra-indication for MASLD (Figure 2A). PPARG is a nuclear receptor that regulates fatty acid and glucose metabolism. In the setting of type 2 diabetes (T2D), active PPARG increases insulin sensitivity by enhancing healthy adipogenesis and increasing production of adiponectin and GLUT4.^49^ As such, PPARG agonists like TZDs are standard in T2D therapy. There are over 50 experimental structures of PPARG with small molecules bound to the same binding pocket as pioglitazone. We docked each compound to PPARG to compare the binding energetics by Rosetta’s Interface Delta X, finding that pioglitazone has superior computational binding energetics to nearly all other docked compounds (Figure 2B). The PPARG-pioglitazone cocrystal structure demonstrates that pioglitazone engages H323, Y473, and H449 with its 2,4-thiazolidinedione ring, which experimentally has been demonstrated (Figure 2C-D).^50^ Its central benzene ring and pyridine tail make several contacts with the hydrophobic interior of the pocket.

**Figure 2.**
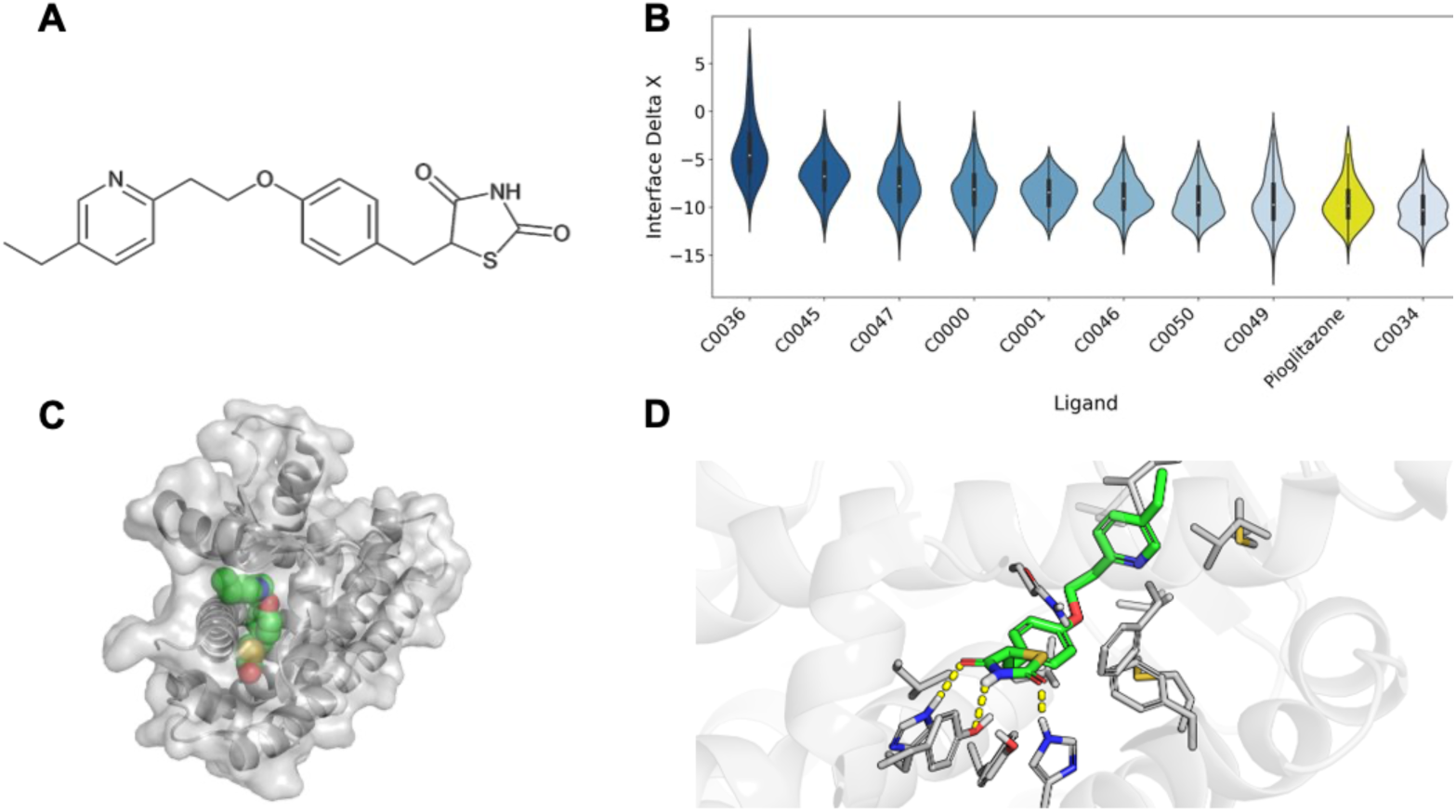
PPARG. Computational modeling of pioglitazone-PPARG interactions. **A)** Chemical structure of pioglitazone. **B)** A computational evaluation of the binding favorability of PPAG with pioglitazone and other small molecules with experimentally resolved structures. Molecules were docked with RosettaLigand and scored by Interface Delta X. **C)** A global view of pioglitazone bound to PPARG. **D)** Specific details of the binding mode between PPARG and pioglitazone, with hydrogen bonds noted in yellow dashed lines.

#### FADS1

FADS1 and FADS2 are integral membrane proteins in the endoplasmic reticulum that catalyze the synthesis of unsaturated fatty acids. FADS1 binds fatty acids in a hydrophobic binding pocket and catalyzes their desaturation via a conserved histidine-rich motif. In our study, changes in FADS1/2/3 expression were used to proxy the effect of icosapent ethyl (Figure 3A) on MASLD risk. However, no experimental structures exist for the FADS1/2/3 proteins to explore the interactions between the drug and protein. To create a computational model for FADS1 bound to icosapent ethyl (Figure 3B-C), we generated an AlphaFold3 model of FADS1 bound to an unsaturated phospholipid. We confirmed this binding pocket and orientation by docking icosapent ethyl to an apo-state FADS1 model using DiffDock, a deep-generative blind ligand docking algorithm. Finally, we docked icosapent ethyl to this FADS1 binding pocket in Rosetta (Figure 3B-C). This model reveals that the lipid tail of icosapent ethyl engages several phenylanalyl and tyrosyl residues in the hydrophobic pocket, and that its hydrophilic ester head group forms a hydrogen bond to ASN (Figure 3C).

**Figure 3.**
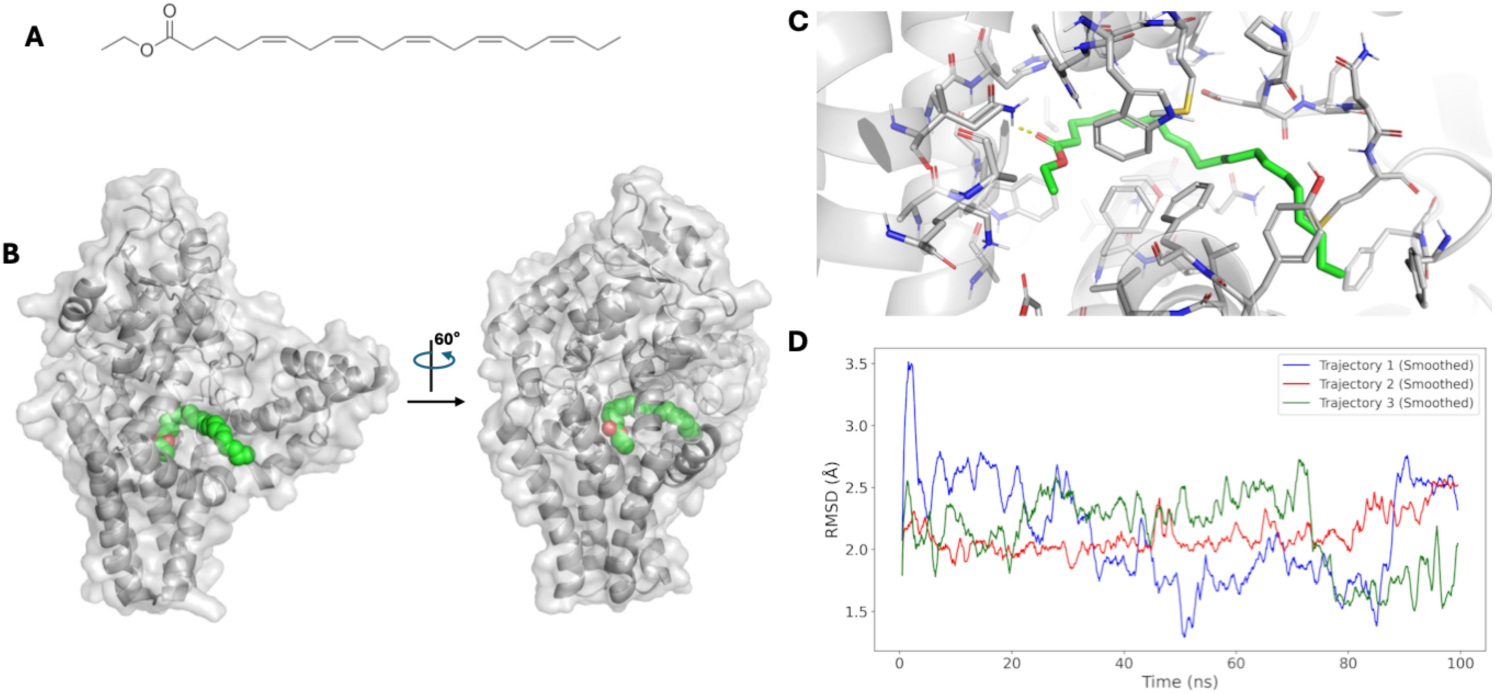
FADS1. Binding mode of IPE to FADS1. **A)** Chemical structure of icosapent ethyl. **B)** Global view of the binding mode of icosapent ethyl (magenta) to FADS1 (gray) and FADS2 (blue). **C)** Comparison of IDX values for 600 docking runs of icosapent ethyl to FADS1 and FADS2. **D)** Binding pocket view demonstrating high sequence and structural homology between FADS1 (gray, transparent) and FADS2 (blue), as well as a similar identified high-scoring binding mode with IPE.

To assess the stability of these interactions, we performed three independent, 100ns unbiased molecular dynamics simulations of the FADS1-icosapent ethyl complex embedded in a lipid bilayer similar in composition to the endoplasmic reticulum (60% POPC, 30% POPE, 5% POPS, 5% cholesterol). Icosapent ethyl remains bound to FADS1 throughout each trajectory (Supplemental Movie 1). These trajectories demonstrate that hydrophilic interactions with icosapent ethyl’s head are stable, but that flexibility exists in the hydrophobic tail region, resulting in a 1.5-2.75Å root-mean square deviation (RMSD) of icosapent ethyl to its initial Rosetta-identified binding pose over the duration of each simulation (Figure 3D). Together, these models suggest a binding mode for icosapent ethyl to FADS1 with the flexibility expected of a long alkyl chain. Finally, we constructed a model of FADS2 bound to icosapent ethyl via the same procedure. Due to the nearly identical sequence and structural homology of the FADS1 and FADS2 binding pockets, MD simulations were not run for the FADS2-icosapent ethyl complex complex (Figure 4A-C). Indeed, structural models made with RosettaLigand demonstrate nearly identical binding of icosapent ethyl to both FADS1 and FADS2 (average Interface Delta X –14.199 vs –14.058, respectively).

**Figure 4.**
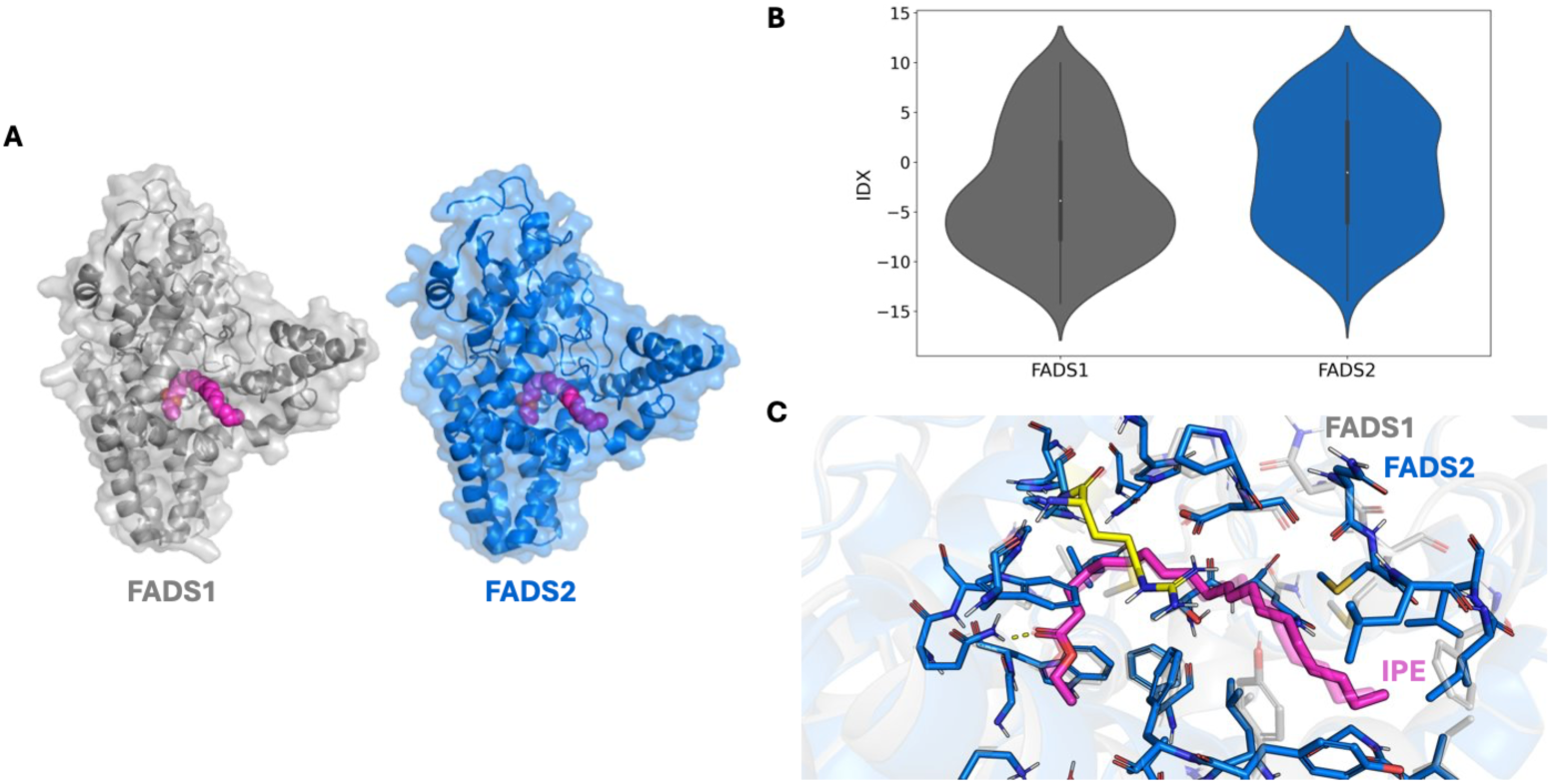
FADS2. Binding mode of icosapent ethyl to FADS2. **A)** Global view of the binding mode of icosapent ethyl (magenta) to FADS1 (gray) and FADS2 (blue) **B)** Comparison of IDX values for 600 docking runs of IPE to FADS1 and FADS2. **C)** Binding pocket view demonstrating high sequence and structural homology between FADS1 (gray, transparent) and FADS2 (blue), as well as a similar identified high-scoring binding mode with icosapent ethyl.

#### S1PR2

Sphingosine-1-phosphate (S1P) is a bioactive lipid that binds to S1P receptor subtypes (S1PR1-5) to mediate cellular activities of the immune, cardiovascular and nervous systems.^51^ S1PR2 is an emerging drug target in multiple sclerosis^52^, which was the primary indication used for fingolimod in the MR, due to its implication in CNS demyelination. Fingolimod has structural similarity to S1P (Figure 5A) and known efficacy through binding S1PR1. Docking fingolimod into an experimental structure of S1PR2 bound to S1P (PDB: 7T6B) with Rosetta reveals that fingolimod’s putative binding mode to S1PR2 is highly homologous to that of S1P (Figure 5B-F). Both engage the hydrophobic core of S1PR2 with unsaturated alkyl chains, and both form hydrogen bonds to S1PR2 with hydrophilic head groups (Figure 5C-F). However, S1P’s hydrophilic head group is larger, allowing it to form meaningful hydrogen bonds to S1PR2 that fingolimod cannot access (Figure 5C-F).

**Figure 5.**
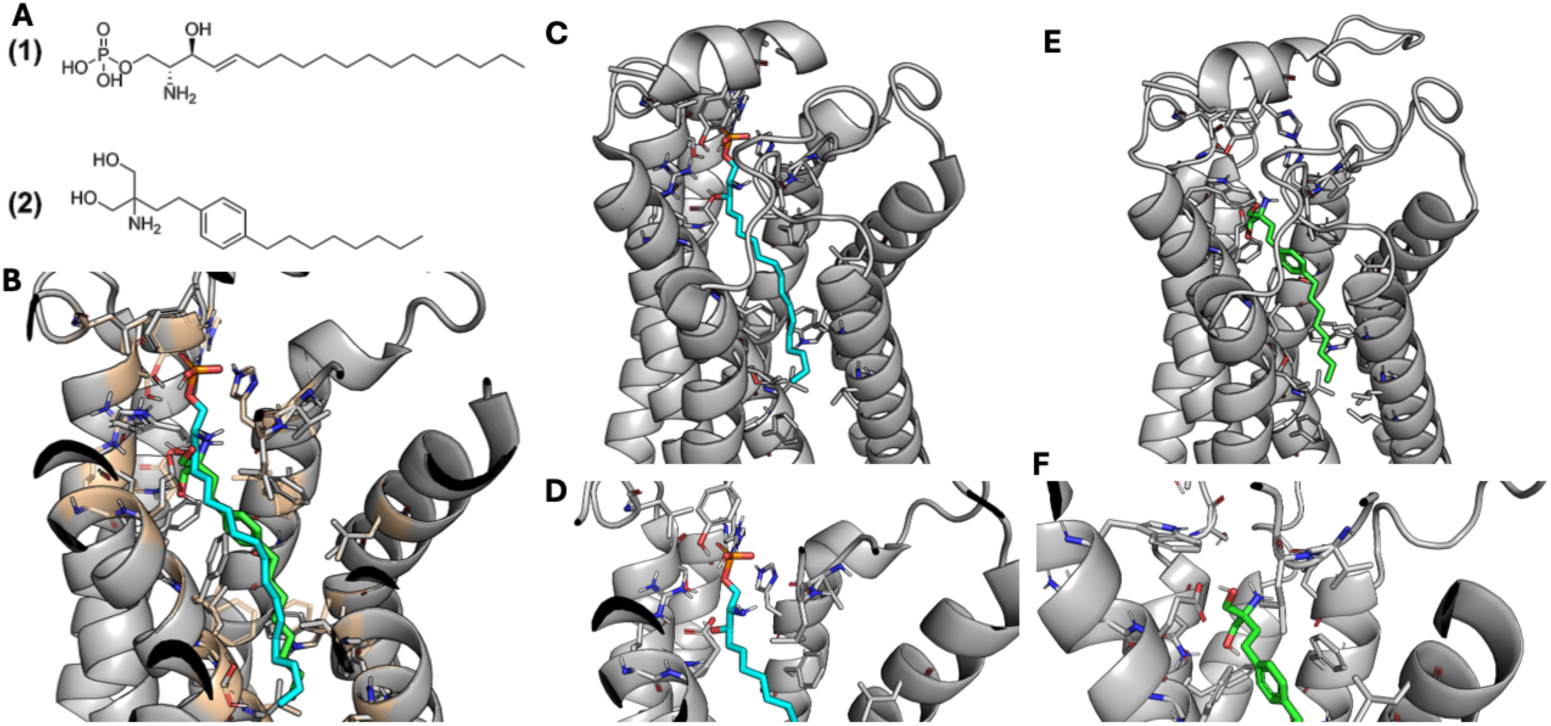
S1PR2. Computational modeling of fingolimod-S1PR2 interactions. **A)** Chemical structure of sphingosine-1-phospage (S1P) (1) and fingolimod (2). **B)** Global comparison of the binding modes of S1P (cyan) and fingolimod (green) to S1PR2 identified by RosettaLigand. **C-F)** Detailed views of binding mode of S1PR2 to S1P (**C-D**) and fingolimod (**E-F**).

## Discussion

Although new medications are beginning to emerge, relatively few treatment options exist for MASLD. Therefore, finding ways to repurpose existing medications remains crucial for disease management for the ∼84 million Americans living with MASLD^14^. This study identified icosapent ethyl and fingolimod as potential treatments for MASLD through integration of genetic analyses, protein dynamics, and pathway analysis. In addition, our MR analysis indicates that increasing *PPARG* expression increases risk of MASLD, which conflicts with data of pioglitazone hydrochloride from clinical trials.^53^

We adapted a previously developed genetically informed drug-repurposing pipeline^34^ for MASLD management. We identified drug-gene pairs by mapping gene expression signatures from the largest GWAS study on MASLD to existing drug targets using publicly available databases. We validated these drug-gene pairs by predicting changes in MASLD risk using the MR framework. Further evidence was derived through protein folding/molecular dynamics and pathway analyses, which involved creating the first computational model of FADS1 bound to icosapent ethyl, to our knowledge.

Our analysis revealed that mapping based on directional genetic effects did not eliminate all potential drug targets that may exacerbate liver disease, as some targets identified are known to increase the risk of drug-induced liver injury, as reported in the NCBI LiverTox ^38^ or FDA DILIRank.^39^ We anticipated that filtering based on the direction of effect using GPGE —which integrates both the direction of genetic effects and drug mechanisms to inform pharmacological modulation—would help identify drug targets more likely to treat MASLD rather than exacerbate it. Proxying drug effects may be difficult using gene expression, given drugs typically act on proteins rather than gene expression itself. However, our choice to select increased *FADS1* expression to proxy icosapent ethyl, a FADS1 agonist and synthetic derivative of the omega-3 fatty acid eicospentaenoic acid (EPA), was supported by a previously study which showed that increased *FADS1* expression was associated with the largest increase in circulating EPA levels compared to other fatty acid metabolism genes. ^21^

Our findings align with results from a large-scale phenome-wide drug repurposing screen^54^, which demonstrated that limiting drug targets to those with directional genetic evidence did not improve the proportion of successful drug indications as they advanced through clinical trials. It should be noted that we selected drug targets based on their association with gene expression in healthy, disease-relevant tissue rather than with clinical pathology. Using gene expression from healthy tissue may explain discrepancies in filtering results based on the direction of effect. To address this gap, future studies should integrate transcriptomic data from diseased tissues to support the translational relevance of genetic findings and refine drug target identification strategies.

Our observations suggest that the activation of *FADS1* expression, as a proxy for icosapent ethyl treatment, confers a protective effect against MASLD, which can be attributed to several biochemical and metabolic mechanisms. FADS1, or fatty acid desaturase 1, is an integral membrane protein that catalyzes the synthesis of unsaturated fatty acids. Icosapent ethyl functions by lowering triglyceride levels through the dual mechanisms of decreasing synthesis and enhancing clearance.^55^ Traditionally, icosapent ethyl is used to treat patients with hypertriglyceridemia and to reduce cardiovascular events. The Reduction of Cardiovascular Events with icosapent ethyl-Intervention Trial (REDUCE-IT) demonstrated that icosapent ethyl significantly reduced both triglyceride levels and ischemic events in statin-treated patients with elevated triglycerides and either cardiovascular disease (CVD) or diabetes.^56,57^ Given that MASLD and CVD share multiple risk factors, including obesity, type 2 diabetes, and insulin-resistance^58^, and that hypertriglyceridemia is a well-established contributor to MASLD development, the lipid lowering properties of icosapent ethyl may hold broader implications for MASLD treatment. However, the effects of this trial may be incomplete,^59,60^ suggesting the need for follow-up studies to clarify and confirm the protective effects of icosapent ethyl. Finally, enhanced FADS1 activity leads to an increased conversion of arachidonic acid to EPA, which is less pro-inflammatory than products derived from arachidonic acid metabolism. This reduces the inflammatory response and ameliorates lipid dysregulation central to MASLD development.^61^ Our findings also identified lipid metabolism as one of the most significantly implicated pathways in the pathway analysis. Further, MR analysis predicted a significant reduction in MASLD risk with proxied icosapent ethyl treatment. Together, these data suggest that icosapent ethyl has strong potential for repurposing in MASLD management, by improving lipid control and potentially reducing liver enzyme levels.

Our findings indicate that increases in *S1PR2* expression, proxying the effects of fingolimod, exerts a protective effect against MASLD. S1PR2 is a cell surface receptor that plays a significant role in lipid metabolism and immune response. Fingolimod, originally developed as an immunomodulatory drug for multiple sclerosis, is known to interact with S1P receptors. Fingolimod, an S1PR modulator, has structural similarity to S1P with low affinity for S1PR2. Studies have found it is capable of modulating S1P signaling pathways involved in maintaining lipid homeostasis (e.g. regulation of lipogenesis, fatty acid oxidation, and lipid transport) and, as such, may prevent lipid accumulation in the liver^62^. The activation of S1PR2 by fingolimod may also contribute to improved insulin sensitivity^63^ and reduced hepatic inflammation.^64^ As far as sphingolipid metabolism is concerned, there have been no clinical studies yet, but fingolimod and SK2 inhibitor K145276 have ameliorated NAFLD in mouse models.^65^ *S1PR2*, which showed strong genetic signals in this study, is highly expressed in vascular tissue and smooth muscle, including liver, heart, kidney, lung, and brain.^66^ Interestingly, *S1PR2* was the only gene with a strong association in liver tissue in our GPGE analyses. Downstream signaling pathways initiated by *S1PR2* also play a role in multiple sclerosis, fibrosis, and inflammation.^67^ Our protein docking analyses revealed that fingolimod’s binding mode to S1PR2 is highly homologous to that of S1P. One pathological change associated with MASLD includes liver cell damage caused by inflammation. Our data is consistent with a model where fingolimod treats MASLD through regulation of lipid metabolism and inflammation regulation. Ligand analysis revealed there are limited targets currently available. However, our results suggest that S1PR2 could be a promising drug target for future mechanistic drug discovery studies.

In our study, we observed an increased risk of MASLD associated with increased *PPARG* expression, a finding that contrasts with the beneficial effects of PPARγ agonists in managing type 2 diabetes and observations from clinical trial data suggesting MASH resolution in individuals with type 2 diabetes.^68–70^ A recent meta-analysis of the PPARγ agonist, pioglitazone which is a TZS, in individuals with MASLD, prediabetes, and type 2 diabetes reported improvement in steatosis and in resolution of steatohepatitis.^71^ It is likely that such a strategy of targeting PPARγ has not been pursued more aggressively because of the adverse effect profile of TZDs, most notably oedema, fluid retention, and heart failure. These results are reflected in recent clinical practice guideline recommendations from the European Association for the Study of the Liver, European Association for the Study of Diabetes, and European Association for the Study of Obesity (EASL/EASD/EASO) that assign pioglitazone a weak treatment recommendation and not recommended as MASH-targeted therapy.^72^

Our genetic study suggests that increased expression of basal *PPARG* levels is associated with increased risk of MASLD. Peroxisome proliferator-activated receptors, including PPARG, are nuclear receptors that form heterodimers with retinoid X receptors (RXR) and act as transcription factors. Their role in transcriptional regulation can lead to variable and, at times, paradoxical effects, particularly during adipogenesis. Suggesting differential activation and cell-specific effects of PPARG (pharmacological versus intrinsic activation) may explain these findings. Mouse models with *Pparg* selectively knocked out from hepatocytes demonstrate that PPARγ acts as a steatogenic factor in these cells.^50,73^ Similarly, mice overexpressing PPARγ exhibit increased expression of steatogenic genes.^74^ Treatment with rosiglitazone or pioglitazone in mice that naturally express PPARγ leads to an increase in liver steatosis^75–77^ ^78^ when fed high fat diets. In overweight mice, however, pioglitazone decreases hepatic fat accumulation.^79^ Consequently, enhanced expression of PPARγ in the liver, likely resulting from increased fat accumulation in hepatocytes, may diminish the therapeutic effects of TZDs on liver steatosis, which are primarily due to their insulin-sensitizing actions in peripheral tissues. Therefore, the specific activation of PPARγ in hepatocytes versus non-parenchymal hepatic cells might explain discrepancies in efficacy between genetic predispositions and pharmacological impacts on steatosis in clinical settings. Further studies are needed to assess whether a genetic predisposition involving PPARγ might reduce or limit the effectiveness of TZD treatments for NAFLD, as observed in mice.^78^

The GWAS utilized in this study was based on liver enzyme levels rather than MASLD diagnosis which could introduce the possibility that some signals may be related to enzyme kinetics or systemic perturbations rather than the disease itself. However, we replicated 81 genes identified in a histologic cohort in which MASLD was defined using liver biopsies and an intendent radiologic cohort where MASLD was determined using CT/MRI imaging data.^26^ These phenotype definitions for MASLD are considered the most comprehensive. All significant MR genes were replicated in both the radiological imaging and liver biopsy data, which provides strong evidence for the reliability of the genetic signals detected in this study.

The absence of certain expected positive controls, such as FGF21 and GLP1R, in our findings is primarily due to our analytical framework, which prioritized only putative effector genes based on a large MASLD GWAS where these genes were not associated with MASLD susceptibility. However, while THRB, another potential positive control, was not directly identified in our gene-based expression analysis, it is a nuclear receptor that binds with RXRs, which were among the strongest signals in our analyses. This suggests that THRB-targeting may not be substantially more specific than targeting other RXR binding partners and the lack of direct THRB signal is likely an artifact of restricting causal discovery to TWAS-significant genes. Expanding the analysis to include all TWAS results may reveal additional pathway-level insights, but we anticipate that RXR- and diabetes-related signaling would remain prominent and that the significance threshold adds rigor to the analytical framework.

As with any MR analysis, the genetic instruments used in this study have weaknesses, which should be considered when interpreting results (Supplemental Table 1). However, the strength of MR results can be enhanced by incorporating additional supportive information, as we have done in this study. For example, the use of computational tools such as Rosetta and DiffDock add depth to the mechanistic understanding of the proposed drug-gene interactions, supporting the rationale for their repurposing for MASLD treatment. Like MR, interpretation of TWAS results should be carefully considered. For example, GPGE is designed to model the basal state of a population and may not account for system perturbations. However, the comprehensive design of this study provides additional lines of evidence. Further validation of our findings could be pursued using methods such as self-controlled case series.^34^

In summary, our study demonstrates that contextualizing genetic signals through pathway analysis and protein folding experiments can elucidate potential drug repurposing targets and novel therapeutic discoveries for MASLD. We have incorporated protein dynamics into our analysis, adding an additional layer of knowledge and interpretation of mechanisms involved in MASLD. While the presence of a genetic effect can indicate system-wide changes, these signals may have limited meaning without understanding the impact on protein function. By integrating these new dimensions, we provide a more nuanced and comprehensive understanding of potential therapeutic targets that can be leveraged for MASLD treatment. Overall, these analyses integrated genetic, molecular, and pharmacological data to further advance our understanding of MASLD, elucidating both novel genetic associations and potential therapeutic interventions.

## Data Availability

The GWAS data used in this study are available through dbGAP under accession number phs001672.v7.p1 (Veterans Administration MVP Summary Results from Omics Studies). All code is available upon request.

https://www.ncbi.nlm.nih.gov/projects/gap/cgi-bin/study.cgi?study_id=phs001672.v7.p1

## Acknowledgements

We gratefully acknowledge the Veterans who participated in the Million Veteran Program. This research is based on data from the Million Veteran Program, Office of Research and Development, and Veterans Health Administration (MVP003/028). This publication does not represent the views of the Department of Veterans Affairs or the United States Government. Support provided by T32GM145734-01 (HMS), BX003362 (Chang/Tsao), K12AR084232 (JNH & MMS), VA BLR&D IK2-BX006551 and Doris Duke Foundation 2023-0224 (MGL), CSP2012 (KB, KL, JAL), T32HL00773 (ATA), and R01DK134575 (MV). JAL, KL, and MV are also supported by MVP003.

## Author contributions

HMS, ATA, JAD, JNH, JHB, JK, ML, and SK performed the analyses. HMS drafted the initial manuscript, which was reviewed and edited by MV, TLE, MMS, NKK, ML, DG, and KMC. MV, TLE, MMS, NKK, and JAD led the primary study design, with contributions from AB, KL, DG, JSL, KH, DRM, and PR.

## Web resources

DgIDB: https://dgidb.org

OpenTargets: https://www.opentargets.org

DrugBank: https://go.drugbank.com/

FDA DILIRank: https://www.fda.gov/science-research/liver-toxicity-knowledge-base-ltkb/drug-induced-liver-injury-rank-dilirank-dataset

NCBI LiverTox: https://www.ncbi.nlm.nih.gov/books/NBK547852/

GTEx: https://gtexportal.org/home/

DECODE: https://download.decode.is/form/folder/proteomics

AlphaFold: https://alphafold.ebi.ac.uk/

## Data and code availability

The GWAS data used in this study are available through dbGAP under accession number <u>phs001672.v7.p1</u> (Veterans Administration MVP Summary Results from Omics Studies). All code is available upon request.

